# Infection rate of *Borrelia burgdorferi* genospecies in human-biting *Ixodes ricinus* ticks: models for surveillance based on the French citizen science programme CiTIQUE

**DOI:** 10.1101/2025.09.18.25336063

**Authors:** Thierno Madiou Bah, Jonas Durand, Arnaud Cougoul, William Wint, Francesca Dagostin, Thomas Opitz, Xavier Bailly, Pascale Frey-Klett, Karine Chalvet-Monfray

**Affiliations:** University of Clermont Auvergne, INRAE, VetAgro Sup, UMR EPIA, Saint-Genès Champanelle, France; University of Lyon, INRAE, VetAgro Sup, UMR EPIA, Marcy l’Etoile, France; Université de Lorraine, INRAE, IAM, Laboratoire Tous Chercheurs, F-54000 Nancy, France; Environmental Research Group Oxford Ltd, Oxford, United Kingdom; Research and Innovation Centre, Fondazione Edmund Mach, San Michele all’Adige (TN), Italy; Biostatistics and Spatial Processes (UR 546), INRAE, Avignon, France

## Abstract

In Europe, Lyme borreliosis is the most common vector-borne human disease, caused mainly by *Borrelia afzelii and Borrelia garinii*, two species of the *Borrelia burgdorferi* sensu lato (Bbsl) complex transmitted by the tick *Ixodes ricinus*. Accurately assessing the spatial risk of human exposure to these pathogens is essential for efficient public health surveillance. However conventional monitoring often struggles to produce geographically explicit, large-scale data that capture the heterogeneity of human exposure and its drivers. Focusing on continental France, we leveraged data from the French CiTIQUE citizen science programme to analyse spatial variation of Bbsl infection in georeferenced human-biting I. ricinus ticks and to model the relationship between Bbsl distribution and environmental, ecological, and anthropogenic factors. From 2017–2019, 1,891 ticks were analysed, of which 15% tested positive for Bbsl. The most prevalent genospecies were *B. afzelii* (7.2%) and *B. garinii* (4.2%). Infection rates varied spatially, with distinct distribution patterns across pathogen groups. Tick habitat suitability was the most consistent predictor for overall Bbsl infection probability, genospecies-specific models revealed the importance of their respective reservoir hosts: *B. afzelii* occurrence was positively associated with rodent species richness, whereas *B. garinii* was associated with Turdidae species and showed potential traces of a dilution effect due to rodents. Our findings demonstrate the value of citizen science for complementing formal surveillance and provide the first geographically explicit, large-scale insights into Bbsl eco-epidemiology in France. This scalable approach offers an adaptable framework for monitoring vector-borne disease risk and guiding public health strategies.

**Importance:** Lyme borreliosis, caused by *Borrelia burgdorferi* sensu lato, is the most common human vector-borne disease in Europe. Accurate assessment of spatial exposure risk is essential for effective public health surveillance and interventions. Using data from the French CiTIQUE citizen science program, we reveal pathogen-specific spatial patterns and identify the factors shaping them, at a geographic resolution not previously studied. Our findings demonstrate that citizen science can provide a scalable and adaptive framework for long-term surveillance of vector-borne disease risk, offering valuable insights to guide targeted prevention and control measures.

## Introduction

Lyme borreliosis (Lb) is the most prevalent vector-borne human disease in Europe, with approximately 129,000 cases reported annually (1). Though incidence is likely underestimated as Lb is not notifiable in many countries. In France, 39,000 cases were reported in 2023, but recent analyses suggest that the actual incidence may have been closer to 191,000 cases during the same year (2).

Lb is caused by pathogenic spirochetes of the *Borrelia burgdorferi* sensu lato (Bbsl) species complex, primarily transmitted by the hard tick *Ixodes ricinus* (3, 4). In Europe, *Borrelia afzelii* and *Borrelia garinii* are the most common genospecies (5), predominantly maintained by rodents and birds, respectively (6). Their distinct eco-epidemiological cycles, combined with environmental and anthropogenic factors drive spatial heterogeneity in Bbsl distribution (7). Since genospecies differ in clinical outcomes, *B. garinii* being more often linked to neuroborreliosis (8), mapping this variation is critical. Human risk likewise varies widely across landscapes, underscoring the need to map this heterogeneity for surveillance, prevention, and public health action.

Assessing spatial Lb risk typically relies on two layers of information : acarological hazard, defined by the density of host-seeking Bbsl-infected ticks, and human exposure, often approximated by incidence data (5, 9). Both are essential for modelling the local states of the host–vector–pathogen system and predicting infection risk (10). Two complementary approaches are used: mechanistic models, which explicitly represent life-cycle and transmission processes (11), and statistical models (12), which link infection patterns to environmental or socio-economic factors (10, 13).

Field methods, such as host sampling, provide insights into pathogen ecology but rarely capture the full diversity of reservoirs within a single study (e.g., Rataud et al., 2022 for birds in France). Standardised questing tick sampling offers a good proxy for tick density and pathogen hazard (see for instance Wongnak et al., 2022 and Perez et al., 2023). However, tick sampling is often not standardized at country level, is limited to small areas or accessible environments and lacks information on human exposure. Human case reports from medical practitioners can provide broader spatial coverage of human risk of infection but are often limited by coarse spatial resolution (17).

To overcome these limitations, citizen science initiatives have emerged as promising tools to generate geographically informed, large-scale data on human-tick encounters. Data that would otherwise be extremely difficult,if not impossible, to obtain through conventional research means, while also fostering engagement and raising awareness (18). In particular, the collection of human-biting ticks is directly associated to human exposure (19), and can be used to study spatial variations in the probability of Bbsl genospecies infection in human-biting ticks.

France hosts diverse climates and environments (20) that support both tick populations and pathogen reservoirs (15, 16, 21). Previous studies on Bbsl distribution drivers in France were conducted at highly localised scales (22, 23), making extrapolation to the contiguous national level difficult. At the other end of the spectrum, European-scale models of Bbsl distribution have been developed (7), but these rely on literature data which are from France limited and at a very local scale, highlighting the need for extensive data and studies to better understand and predict Bbsl risk in this country suffering high Lyme borreliosis incidence (24).

Here, we used human-biting ticks collected through the CiTIQUE citizen-science programme between 2017 and 2019, to study the spatial distribution of Bbsl and its major genospecies across con-tinental France. Combining statistical relative risk mapping, which quantifies the relative density of pathogen presence in relation to its absence, and generalised additive models (GAMs), we characterised the spatial heterogeneity of tick infection risks and identified environmental and ecological drivers shaping the distribution of Bbsl in human-biting ticks, providing a detailed, data-driven picture of Lb eco-epidemiology across continental France.

## Materials and methods

### Tick acquisition

Ticks were collected through the CiTIQUE citizen science programme(www.citique.fr), launched in July 2017 as a collaboration between INRAE (French National Research Institute for Agriculture, Food and Environment), the Laboratory of Excellence ARBRE, Anses (French Agency for Food, Environmental and Occupational Health & Safety), and the CPIE network (Permanent Centre for Environmental Initiatives). This programme aims to improve the understanding of the ecology of ticks and the tick-borne diseases they transmit, particularly Lyme Borreliosis, in order to support prevention efforts based on monitoring.

Citizens can participate in the CiTIQUE programme choosing among various levels of involvement, ranging from promoting the programme to actively contributing to research through activities in the open lab “Tous Chercheurs”, or by reporting tick bites and collecting and sending biting ticks to the tick bank maintained by the national programme. Tick bites can be reported via website, mobile application, or paper form, with information on date, GPS location, basic personal data (age, sex, activity, bite localisation), and environmental context (e.g., forest, garden, meadow). These data are linked to the tick samples upon reception. Between 2017 and 2020, more than 17,000 human tick-bite reports were submitted, with over 4,500 of these associated with one or more tick specimens.

### Pathogen identifications

A total of 2009 ticks, collected between 2017 to 2019 (inclusive), were randomly selected for DNA extraction, with the objective of including at least 150 human-biting ticks for each French NUTS-1 regions (Nomenclature of Territorial Units for Statistics - major socio-economic regions) except for Provence-Alpes-Côte d’Azur (PAC) with 59 records. From this dataset, 1 891 *Ixodes ricinus* ticks were retained for the modelling analyses presented in this study, including 221 Adults, 1324 Nymphs and 110 Larvae, with 236 undetermined specimens. Details on the overall sample, selection procedure, and dataset structure can be found in (25).

The tick stages and species were identified morphologically using identification keys by Pérez-Eid (2007) and Estrada-Pena et al. (2018). Species identification was confirmed using specific primers and probes on the microfluidic real-time PCR assay described below. Tick DNA was extracted and screened for pathogens using the method described in Melis et al. (2024). Briefly, tick DNA was extracted using NucleoSpin® Tissue kit (Macherey-Nagel, Germany), following manufacturer’s instructions. Then, all samples underwent a pre-amplification step by PCR with the Preamp Master Mix (Standard BioTools, USA). High-throughput microfluidic real-time PCR was then performed on a BioMark™ real-time PCR system (Standard BioTools, USA), using the 48.48 Dynamic Array™ (Standard BioTools, USA) to detect pathogens and identify ticks. Only results for the different Bbsl genospecies are presented and used in this paper: *Borrelia afzelii, Borrelia garinii, Borrelia burgdorferi* sensu stricto (s.s.), *Borrelia valaisiana, Borrelia spielmanii, Borrelia bissettii, Borrelia lusitaniae*. This method cannot detect coinfections between different *Borrelia* genospecies. The sequences of the primers and probes are provided in Michelet et al. (2014).

### Covariates acquisition

To explain Bbsl spatial distribution, we extracted covariates related to the density of host-seeking *I. ricinus*, pathogen occurrence and persistence, and human exposure. Climatic variables were included for their effect on tick habitat suitability (30), along with habitat suitability indices for *I. ricinus* and host-related variables. Non-competent hosts (e.g., roe deer) were also considered for their role as tick amplifiers (31). Because pathogens were identified in human-biting ticks, we further included variables reflecting human exposure, such as population density and human pressure.

In total, 103 covariates were extracted at a 5-km grid resolution across continental France (Cf. Supplementary table 1). To reduce collinearity and overfitting, covariates were first grouped into seven categories: bioclimatic, land cover/soil, human pressure, and host-related (deer, birds, rodents, species richness). Then, within each category, hierarchical clustering on principal components (HCPC) was applied, with clusters defined by the largest relative loss of inertia. The most relevant covariates within each cluster were selected based on literature and prior hypotheses (Cf. Table 1).

**Table 1:** Covariates selected for the models with their descriptions and associated hypotheses on their effects on Lyme borreliosis occurrence.

| Type | Variable | Description | Resolution | Hypotheses | Source |
| --- | --- | --- | --- | --- | --- |
| Vectors | Habitat suitability index of *I. ricinus* in France | Habitat suitability index for *I. ricinus* through-out continental France multi-criteria decision analysis integrating climate, land cover, altitude and the density of wild ungulates. | 100m | Area suitable for *Ixodes ricinus* may be area with high density of *I. ricinus* and as such Bbsl prevalence as *I. ricinus* is the main vector species for *B. burgdorferi* sensu lato (Bbsl) in France (3, 16) | (21) |
|  | *I. ricinus* suitability | Proportion of suitable habitat for *I. ricinus* | 1km | Area suitable for *I. ricinus* may be area with high density of *I. ricinus* and as such Bbsl prevalence *I. ricinus* is the main vector species for Bbsl in France (3, 16) | William Wint Mood project (undisclosed data) |
| Hosts | Deer | Sum of ensemble models describing the proportion of suitable habitat for roe deer, red deer, and fallow deer respectively within recorded distributions for Europe as identified from diverse source | 1km | High cervid densities are hypothesised to increase overall tick populations without directly increasing Bbsl prevalence, as cervids are incompetent reservoirs. However, their role in supporting adult tick feeding indirectly sustains nymphal tick density, which contributes to Bbsl transmission dynamics in areas with abundant reservoir-competent hosts (33). | William Wint Mood project (undisclosed data) |
|  | Mammal species richness | Mammal species richness including major taxonomic Orders (Cetartiodactyla, Carnivora, Primates, Eulipotyphla, Chiroptera, Rodentia) and marsupials | 10km | High mammal species richness is hypothesised to elevate Bbsl prevalence in ticks by providing multiple competent hosts, which support ongoing infection cycles within tick populations (34). However, in highly diverse communities, the "dilution effect" might reduce overall infection rates by increasing the proportion of non-competent hosts (35). | Biodiversity mapping (36) |
|  | Sheep and cattle density | Density of cattle and sheep per km<sup>2</sup> with weight estimated by Random Forest model with a dasymetric method | ~10 km | Livestock grazing may affect negatively the abundance of small rodents and ticks (37). High livestock densities are correlated with areas of human activity, potentially leading to increased reports of biting ticks | Gridded livestock of the world database (38) |
|  | Index of rodent species richness | Predicted average number of rodent species in the field.<br>Layer containing 5 rodent species: *Apodemus agrarius*, *Apodemus flavicollis*, *Myodes glareolus*, *Microtus arvensis*, *Apodemus sylvaticus* | 1km | Higher rodent species richness is hypothesised to elevate Bbsl. prevalence in ticks by increasing the diversity of reservoir hosts for genospecies such as *B. afzelii*. This enhances local transmission dynamics and may sustain infection cycles across various habitats (39). | (40) |
|  | *Myodes glareolus* suitability map | Ensemble model describing the proportion of suitable habitat for *M. glareolus* | 1km | *Myodes glareolus* is a good reservoir for Bbsl (41) | William Wint Mood project (undisclosed data) |
|  | Anthro bird abundance | Layer containing the sum of the predicted full year abundance of 4 species which habitat extend to anthropised areas: *Sylvia atricapilla*, *Parus major*, *Luscinia megarhynchos*, *Ficedula albicollis* | 3km | Birds that can thrive in anthropised habitat can be more susceptible to carry *B. garinii* (6). | E-birds |
|  | Turdidae abundance | Sum of abundance of birds belonging to the Turdidae family.<br>Layer containing the sum of the predicted mean full year abundance of birds belonging to the Turdidae family: *Turdus iliacus*, *Turdus merula*, *Turdus philomelos*, *Turdus pilaris* | 3km | Members of the Turdidae family are reported to be hosts of *B. garinii* (42)<br>*Turdidae* bird abundance is hypothesised to increase Bbsl prevalence in ticks by serving as key reservoir hosts, particularly for bird-specific *Borrelia* genospecies as *B. garinii*. Their behaviour and high tick burden facilitate *Borrelia* transmission, increasing infection rates in tick populations in areas where these birds are common. | E-birds |
|  | Passeriformes species richness | Passeriformes species richness | 10km | High passeriform species richness may be associated with more *B. garinii* prevalence (43) | Biodiversity mapping (36) |
| Bioclimatic | Mean temperatures of the driest quarter | Average mean temperature for the years 1970-2000 of the driest quarter of the year. A quarter is a period of three months | 1km | An association between temperature and *B. burgdorferi sensu lato* prevalence has been pointed out (7). Mean daily temperatures during the driest quarter might modify the prevalence of *Borrelia burgdorferi* in ticks by impacting tick survival (44), questing activity (15), and pathogen transmission (45). Optimal temperatures would lead to larger tick populations and longer active periods, leading to elevated *Borrelia* transmission. | Worldcilm 2 (46) |
|  | Temperature seasonality | Standard deviation of the monthly mean temperatures for the years 1970-2000 | 1km | Temperature seasonality, especially shifts from colder to warmer periods, is hypothesised to increase Bbsl prevalence in ticks (7), potentially by synchronizing host-seeking behaviour with reservoir host availability (47), extending active seasons, and enabling differential pathogen genotype survival across varying climate (48). | Worldcilm 2 (46) |
|  | Annual precipitation amount | Accumulated precipitation amounts over 1 year for the years 1970-2000 | 1km | Higher annual precipitation is hypothesised to increase *Borrelia burgdorferi sensu lato* prevalence in ticks by supporting optimal moisture conditions and vegetation that sustain tick populations and questing behaviour (49, 50), enhancing pathogen transmission across tick-host networks. | Worldcilm 2 (46) |
|  | Mean precipitation amount of the driest quarter | Average mean precipitation for the years 1970-2000 of the driest quarter of the year. A quarter is a period of three months | 1km | Humidity plays a vital role in tick activity and survival (51). Higher mean monthly precipitation in the driest quarter is hypothesised to increase *Borrelia burgdorferi sensu lato* prevalence by enhancing tick survival and activity during dry periods, thereby supporting sustained transmission cycles of *Borrelia* in regions prone to seasonal drought as in USA (52, 53). | Worldcilm 2 (46) |
| Land cover | Shrubs cover fraction | Fractional cover (%) in a 100m pixel for the shrubland class | 100m | Shrub cover fraction is hypothesised to positively influence *Borrelia burgdorferi sensu lato*, particularly *Borrelia garinii* as shrubs can provide varied resources to birds species in the form of food, nesting and shelter from predators (54). Additionally, shrub habitats can be associated with more species richness and in some cases abundance (55, 56). | Copernicus global land service 2019 (57) |
|  | Deciduous broad leaf forest | (Discrete) forest type of layer for all pixels where the tree cover fraction exceeds 1% and matches the deciduous broad leaf forest | 100m | Deciduous forests might act as critical habitats for maintaining and amplifying *Borrelia* transmission cycles due to favourable microclimatic conditions, high host diversity, and abundant tick populations (58, 59). However, these forests' exact role in amplifying or diluting *Borrelia* prevalence depends on the interplay between biodiversity, host availability, and habitat structure. | Copernicus global land service 2019 (57) |
|  | Grass cover fraction | Fractional cover (%) in a 100m pixel for the herbaceous vegetation class | 100m | Grass cover fraction is hypothesised to influence Bbsl prevalence in ticks by supporting tick habitats in ecotone areas and near woodland edges, though prevalence is generally lower than in forested regions. Grasslands with adjacent habitats for reservoir hosts may contribute to localised *Borrelia* transmission dynamics. | Copernicus global land service 2019 (57) |
|  | Crops cover fraction | Fractional cover (%) in a 100m pixel for the cropland class | 100m | Crop cover fraction is hypothesised to influence Bbsl prevalence in ticks due to the lack of suitable habitats, reservoir host diversity, and management practices in agricultural landscapes. Edge habitats near croplands, however, may support occasional tick populations and pathogen transmission, though at reduced prevalence levels compared to forested areas (60). | Copernicus global land service 2019 (57) |
|  | Built-up cover fraction | Fractional cover (%) in a 100m pixel for the built-up class | 100m | Built-up areas can alter the natural habitat favourable for *I. ricinus* and Bbsl reservoir, though some connected greenspaces in urban environment may support enough wildlife such as rodents and birds to support Bbsl transmission cycle (61–63). | Copernicus global land service 2019 (57) |
| Human pressure | Travel times cities | Accessibility to high-density urban centres in 2015 as measured by pixel-level travel times for the optimal path between each pixel and its nearest city (that is, with the shortest journey time) | 1km | Travel times to cities impact Bbsl prevalence in ticks, with higher prevalence expected in green spaces of rural areas due to greater connectivity and wildlife reservoirs. Ticks in peri-urban areas may maintain *Borrelia* transmission cycles, while urban areas further from rural habitats would exhibit on average lower prevalence due to reduced wildlife host and tick densities (64). | (65) |
|  | Human population density | Gridded human population estimates | 1km | Areas with high human population density may be areas with a higher probability of tick bite reports and thus tick samples than others. | Worldpop |
|  | Human footprint | Annual dynamics of the global human footprint using eight variables that reflect various aspects of human pressures. | 1km | Human footprint is an important predictor of vector borne disease occurrence (66). Higher human footprint index values are hypothesised to lower overall tick densities but can create isolated, fragmented habitats that maintain *Borrelia* prevalence within tick populations. Urban green spaces and edge habitats support localised *Borrelia* transmission due to the continued presence of tick hosts, even in human-modified landscapes. | (67) |
|  | Soil biodiversity threats | The potential rather than the actual level of threat to soil organism using threats proxy (loss of above ground biodiversity, pollution and nutrient overloading, agricultural use, overgrazing, fire risk, soil erosion, land degradation, climate change) | ~10km | Biotic interactions within soil regulate the structure and functioning of aboveground communities and contribute to the delivery of vital ecosystem services (68, 69). As soil-dwelling arthropods, ticks can benefit from soil biota and plants for their survival (70). We hypothesize that areas with high potential of threats to soil organism were less favourable to ticks and their hosts such as of rodents or soil dwelling birds that respectively benefit of the products of a rich soil , including seeds, fruits, and soil invertebrates. | ESDAC (71) |

For each tick, selected covariate values were extracted within a 5 km radius around the GPS coordinates to account for environmental heterogeneity and geolocation imprecision, and the weighted median, accounting for the proportion of each cell covered by the buffer, was retained (Cf. Table 1). All covariates were centred and scaled before analysis. To further limit collinearity between selected covariates, variance inflation factors (VIFs) were assessed, using a cut-off value of 5, to remove collinear covariates (*car* package, version 3.1.3; Fox and Weisberg, 2018).

### Modelling

We used Generalised Additive Models (GAMs) to investigate the variation in Bbsl and Bbsl genospecies distribution, used as the response variable, in relation to the selected set of continuous explanatory covariates (see Table 1), while accounting for the spatial distribution of the observations. The general structure of the models was:

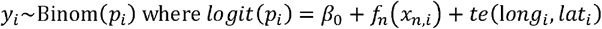

where *f*_*n*_ are spline functions applied to the explanatory covariates *x*_*n,i*_, and *te* is a bivariate tensor product function accounting for the spatial structure based on the longitude and latitude.

Given the large number of 23 explanatory covariates, we applied a double penalty approach to control smoothness of the model terms (i.e., curves *f*_*n*_ and spatial surfaces *te*(long,lat), and perform covariate selection (72). The first penalty controlled the complexity of smooth functions to prevent overfitting by ensuring that relationships remained smooth and interpretable. The second penalty shrunk entire uninformative smooth functions towards zero and effectively removing them from the model. The degree of smoothing was selected using the restricted maximum likelihood (REML) method, and the number of basis functions for smooth terms was limited to the default of 10 simple terms to avoid overfitting.

Three GAM specifications were fitted. The first model (M0) investigated factors associated with the presence of Bbsl across all observations, encompassing both presence (*y*_*i*_ = 1) and absence (*y*_*i*_ = 0) of Bbsl genospecies. In a second step, we focused the modelling process to locations where a *Borrelia* genospecies was detected, to identify factors associated with the infection probability of specific genospecies. Only *B. afzelii* and *B. garinii* had sufficient data for the whole territory, as they were the most frequent genospecies. Therefore, two separate GAMs were constructed (M1 for *B. afzelii* and M2 for *B. garinii*), using the subset of 285 individuals where a Bbsl genospecies was present. In these models, infection with *B. garinii* or *B. afzelii* (depending on the model) was coded as presence (*y*_*i*_ = 1), while infection with another Bbsl genospecies was coded as absence (*y*_*i*_ = 0).

The predicted infection rate of *B. afzelii* and *B. garinii* was calculated by the product of the predicted general bbsl presence (M0) with the species-specific relative probability (M1 or M2 depending on the genospecies).

Model fitting was performed using the *mgcv* package (v1.9.1; Wood, 2011). All final models were checked for residuals validity using the Dharma package. (v0.4.7 ; Hartig et al., 2024).

In addition to GAMs, we used spatial relative risk analysis to identify areas of elevated risk (i.e., “hot spots”) where tick infection by Bbsl and its genospecies was higher than expected, while accounting for underlying tick distribution. Spatial relative risk was estimated using the *sparr* package in R (v2.3.15; Davies et al., 2018). This method estimates the relative density of pathogen presence versus absence points. To account for our spatial sampling, we applied an adaptive bandwidth, allowing finer resolution in densely sampled regions and smoother estimates in sparsely sampled areas. The relative risk surface was estimated asymmetrically, meaning that different bandwidths were applied to the case and control point distributions, as recommended by Davies and Hazelton (2010). Edge effects were corrected using the default settings described by Diggle (1985). Statistical significance of elevated and diminished risk areas was assessed using asymptotic tolerance contours based on p-values, generated via the Monte Carlo method with 1,000 simulations.

All analyses were performed in R version 4.3.2 (78).

## Results

### Descriptive analysis

A total of 1,891 *I. ricinus* ticks were screened for pathogens, of which 291 (15%) were found to be infected with *Borrelia burgdorferi* sensu lato (Bbsl). Among these, *B. afzelii* and *B. garinii* were the most prevalent genospecies, infecting 136 (7.2%) and 80 (4.2%) individual ticks, respectively. Infections with other genospecies were less common, with 37 ticks (2%) infected with *B. valaisiana*, 25 (1.3%) with *B. burgdorferi* ss, 8 (0.4%) with *B. spielmanii*, and 5 (0.3%) with *B. lusitaniae*. No ticks were found to be infected with *B. bissettii*.

*I. ricinus* ticks were collected across the continental French territory, and Bbsl was detected in all NUTS-1 regions. The distribution of infected ticks, however, was uneven: more infected samples relative to the sampling effort were reported in the eastern and central regions, particularly in Grand Est (GES) with the highest infection rate whereas southern regions such as Occitanie (OCC) and Provence-Alpes-Côte d’Azur (PAC) had fewer infected samples (Figure 1).

**Figure 1:**
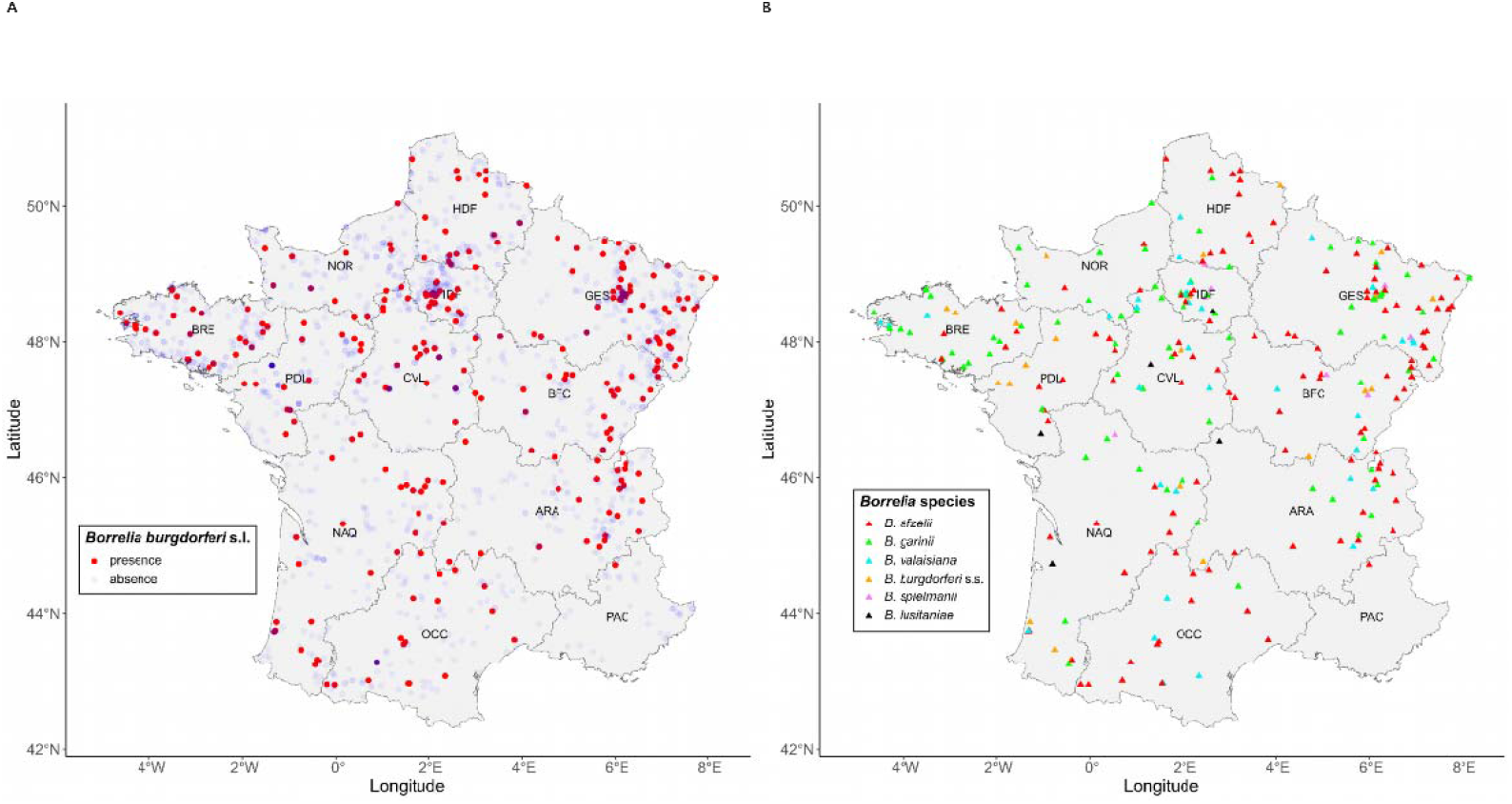
Original location of the 1891 human-biting ticks sent for pathogen identification across the French territory, and associated infection status. Left panel: Red points correspond to sample locations where one *Borrelia* was identified. Blue points mark sample locations where no *Borrelia* was detected. Darker blue shading represents overlapping points due to multiple reports from the same area. Right panel: detailed description of the borrelia genospecies found at each presence location. Each colour indicates the presence of a specific Borrelia burgdorferi genospecies. Major socio-economic (NUTS-1) are as follow : ARA: Auvergne-Rhône-Alpes; BFC: Bourgogne-Franche-Comté; BRE: Bretagne, CVL: Centre-Val de Loire; GES: Grand-Est; HDF: Haut-de-France; IDF: Ile-de-France; NOR: Normandie; NAQ: Nouvelle-Aquitaine; OCC: Occitanie; PDL: Pays-de-la-Loire; PAC: Provence Alpes Côte d’Azur.

### Factors associated with Bbsl distribution

The results of the first GAM model (M0), which investigated factors associated with Bbsl infection rate, are presented in Figure 2. After penalisation, only two variables were retained as significant predictors: the *I. ricinus* habitat suitability index and the grass cover fraction (Table 1). The *I. ricinus* suitability index showed a positive association with Bbsl infection rate (p-value = 0.007), indicating that higher habitat suitability values correspond to an increased probability of Bbsl infection, with this relationship plateauing at higher suitability values (Figure 2D). Bbsl infection rate exhibited a convex relationship with grass cover fraction (p-value = 0.026). At lower grass cover values, an initial increase was associated with a decline in Bbsl infection rate, reaching a minimum, followed by a slight increase at higher grass cover values. However, this latter trend was characterised by greater uncertainty, likely due to the limited number of observations in areas with high grass cover (Figure 2E).

**Figure 2:**
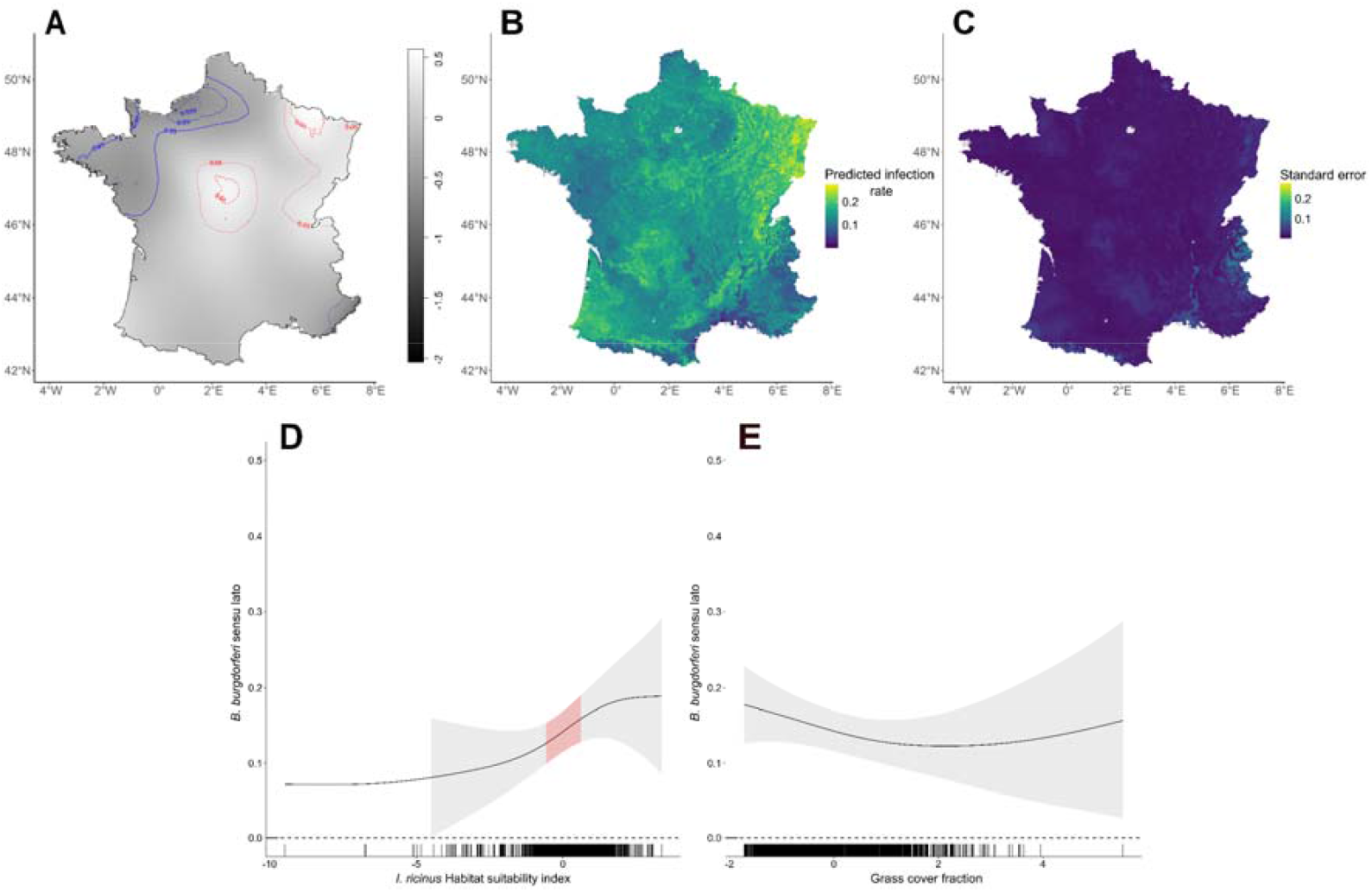
Model predictions for *Borrelia burgdorferi sensu lato* (Bbsl) infection rate in Ixodes ricinus tick across France. A: Bbsl relative risk surface using the Bbsl presence and absence data. Significance contours represent lower risk areas in blue and higher risk areas in red. B: Predicted infection rate of Bbsl based on the M0 GAM, with values ranging from low (dark blue) to high (yellow-green) infection rate. C: Corresponding standard error of the GAM predictions, with lower uncertainty indicated in dark blue and higher uncertainty in yellow. D-E: Each plot demonstrates the marginal effects of *I. ricinus* habitat suitability index (D) and grass cover fraction (E) on Bbsl predicted infection rate, with 95 % confidence intervals shown in shaded areas while values for which the slope is significantly different from zero are highlighted in red. Black ticks along the x-axis represent observed values of the covariables.

The relative risk map (Figure 2A) highlights significant low-risk areas (blue contours), which include the Bretagne (BRE) and Normandie (NOR) regions in northwestern France, and significant high-risk areas (red contours), concentrated in the Grand Est (GES), Bourgogne-Franche-Comté (BFC), and Centre-Val de Loire (CVL) regions in the east and centre of the country. These high-risk regions correspond to areas with higher Bbsl infection rate as predicted by the GAM model (Figure 2B). Conversely, Bretagne (BRE) and Normandie (NOR) exhibited lower predicted infection rates, consistently with the low-risk areas identified by the relative risk analysis. Additional regions, particularly in the southwest and Auvergne-Rhône-Alpes (ARA), also displayed elevated Bbsl infection rates according to the GAM model predictions (Figure 2B). The associated uncertainty in infection rate predictions was highest in the Rhône Valley (southeastern France) and the Alpine regions (eastern France) where sample density was lower and environmental heterogeneity may be greater (Figure 2C).

### Factors associated with Borrelia afzelii distribution

The results of the M1 GAM model assessing factors associated with *B. afzelii* distribution are presented in Figure 3. This model focused on sites where a *Borrelia* genospecies was detected, with the presence probability of *B. afzelii* (conditional on overall *Borrelia* presence) as the response variable. Therefore, the model highlights determinants of the variability in relative incidence of *B. afzelii* among all *Borrelia* occurrences. Four covariates were identified as significant predictors: cattle density, *I. ricinus* habitat suitability index, rodent species richness, and grass cover fraction.

**Figure 3:**
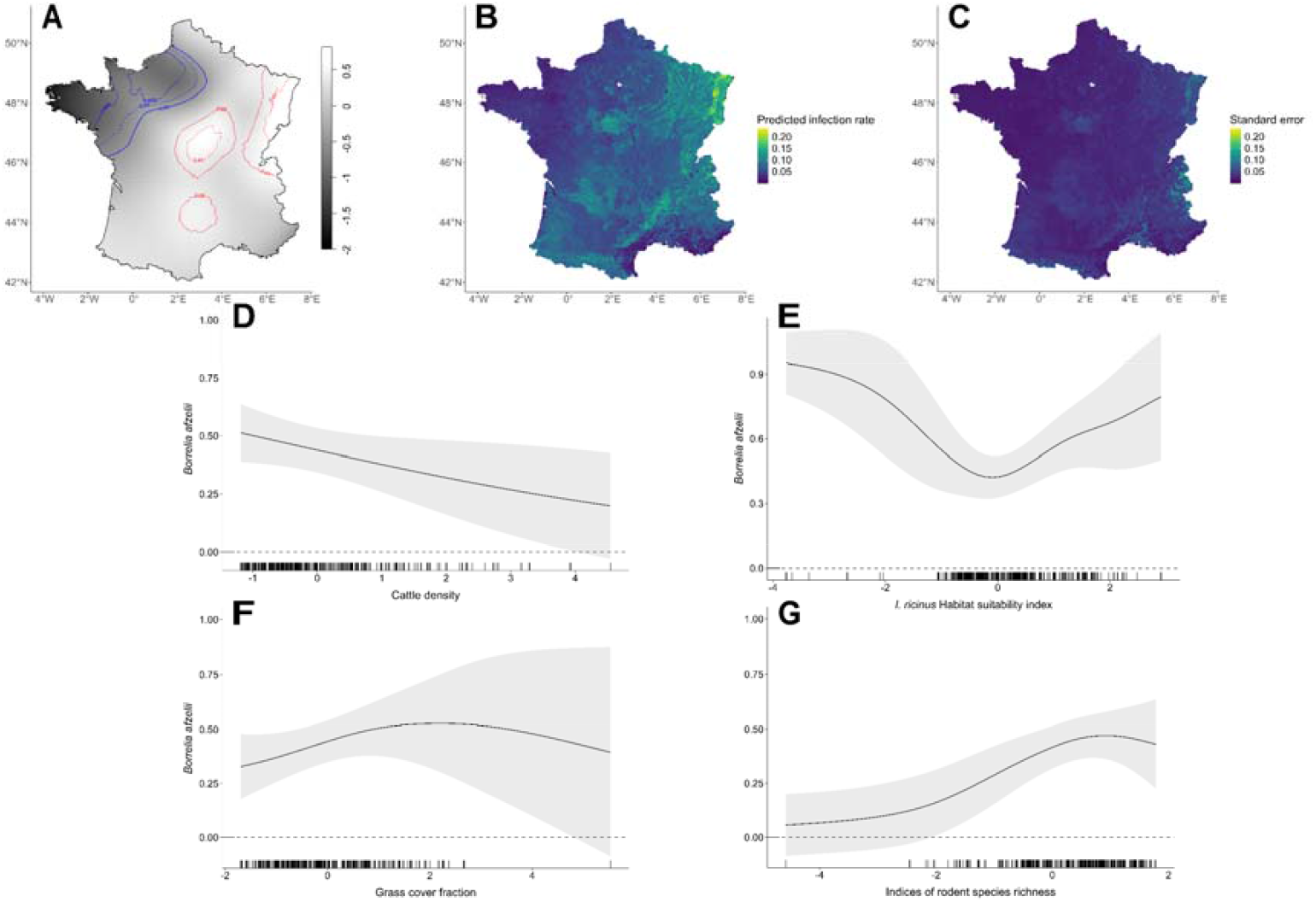
Model predictions for Borrelia afzelii infection rate in *Ixodes ricinus* tick across France. A: *B. afzelii* relative risk surface using the *B. afzelii* presence and absence data . Significance contours represent lower risk areas in blue and higher risk areas in red. B: Predicted infection rate of *B. afzelii* based on the product of the predicted general *Borrelia burgdorferi sensu lato* (Bbsl) presence (M0) with the *B. afzelii* relative probability knowing presence (M1). Infection rate values are ranging from low (dark blue) to high (yellow-green) infection rate. C: Corresponding standard error of the predicted infection rate of *B. afzelii* based on the product of M0 and M1, with lower uncertainty indicated in dark blue and higher uncertainty in yellow. D-G: Each plot demonstrates the marginal effects of the model on the relative probability of having *B. afzelii* knowing Bbsl presence (M1), with Cattle density (D), *I. ricinus* habitat suitability index (E), indices of rodent species richness (F) and grass cover fraction (G). 95 % confidence intervals are shown in shaded areas while values for which the slope is significantly different from zero are highlighted in red. Black ticks along the x-axis represent observed values of the covariables.

Cattle density was negatively associated with *B. afzelii* presence probability (p-value = 0.035), although uncertainty increased at higher cattle densities (Figure 3D). The relationship between B. afze*lii* presence and *I. ricinus* habitat suitability index was convex (p-value =0.009), with a decline in presence probability observed at lower suitability values, reaching a minimum around 0 on the x-axis, followed by an increase at higher suitability values. However, both trends were accompanied by high uncertainty (Figure 3E). A concave relationship was observed with rodent species richness (p-value =0.015), peaking around a richness value of one, before slightly declining. Uncertainty was highest at both low and high richness values (Figure 3F). Grass cover fraction also showed a concave relationship (p-value =0.041), with *B. afzelii* presence probability increasing to a maximum at intermediate grass cover values (around 2 on the x-axis), followed by a slight decline and higher uncertainty at greater grass cover fractions (Figure 3G).

The relative risk map (Figure 3A) revealed that areas of significant low (blue contours) and high (red contours) *B. afzelii* risk broadly overlapped with those identified for Bbsl. However, an additional high-risk area was detected in the northern part of the Occitanie (OCC) region. Predictions from the GAM aligned with the relative risk surface, with the highest *B. afzelii* infection rates in northeastern regions, including GES, BFC and CVL Grand Est, Bourgogne-Franche-Comté, and Centre-Val de Loire. High infection risk was also predicted in regions such as Nouvelle-Aquitaine and Auvergne-Rhône-Alpes (Figure 3B). Prediction uncertainty was greatest in the Rhône Valley (southeastern France), the Alps (eastern France), and the eastern part of the Grand Est region, which also correspond to areas of high predicted infection rates (Figure 3C).

### Factors associated with Borrelia garinii distribution

The predicted infection rate of *B. afzelii* and *B. garinii* was calculated by the product of the predicted general bbsl presence (M0) with the species-specific relative probability (M1 or M2 depending on the genospecies)

The results of the M2 GAM assessing factors associated with *B. garinii* distribution are presented in Figure 4. This model, again restricted to locations where *Borrelia* was detected, used the presence probability of *B. garinii* (conditional on *Borrelia presence*) as the response variable. Therefore, the model highlights determinants of the variability in relative incidence of *B. garinii* among all *Borrelia* occurrences. Two covariates were identified as significant predictors: rodent species richness and *Turdidae* abundance.

**Figure 4:**
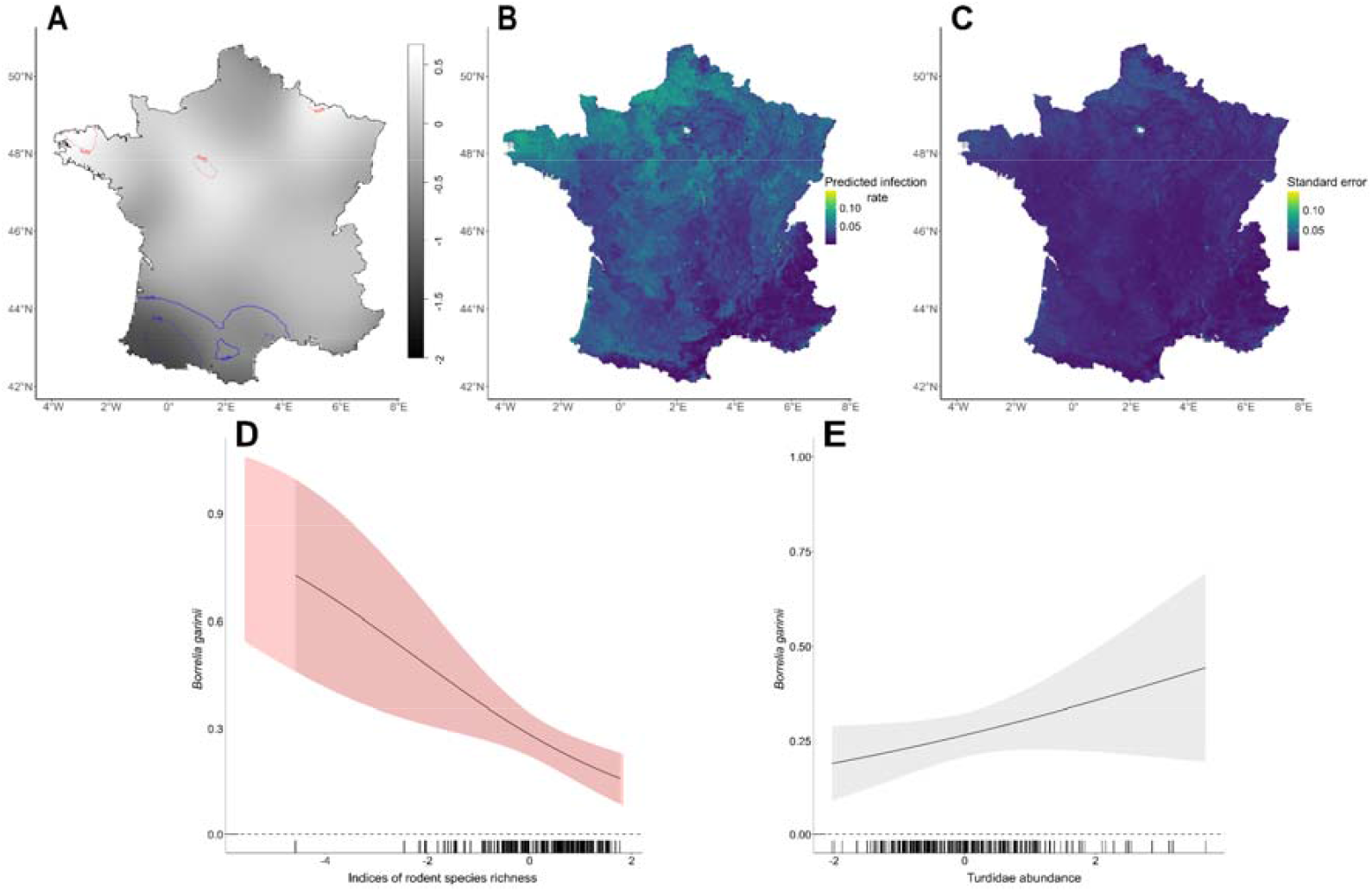
Model predictions for *Borrelia garinii* infection rate in *Ixodes ricinus* tick across France. A: *B. garinii* relative risk surface using the *B. garinii* presence and absence data . Significance contours represent lower risk areas in blue and higher risk areas in red. B: Predicted infection rate of *B. garinii* based on the product of the predicted general *Borrelia burgdorferi sensu lato* (Bbsl) presence (M0) with the *B. garinii* relative probability knowing presence (M2). Infection rate values are ranging from low (dark blue) to high (yellow-green) prevalence. C: Corresponding standard error of the predicted infection rate of B. garinii based on the product of M0 and M2, with lower uncertainty indicated in dark blue and higher uncertainty in yellow. D-F: Each plot shows the marginal effects of the model on the relative probability of having B. garinii knowing Bbsl presence (M2) with indices of rodent species richness (D), *Turdidae abundance* (E) on the probability of having *B. garinii* knowing Bbsl presence. 95 % confidence intervals are shown in shaded areas while values for which the slope is significantly different from zero are highlighted in red. Black ticks along the x-axis represent observed values of the covariables.

Rodent species richness showed a significant negative association with *B. garinii* presence probability (p-value = 0.002), with increased uncertainty at lower values along the x-axis (Figure 4D). In contrast, Turdidae abundance had a significant positive effect on *B. garinii* presence probability (p-value = 0.049), although uncertainty increased markedly at higher abundance values (Figure 4E).

The relative risk map of *B. garinii* (Figure 4A) revealed localised high-risk areas (red contours) in the western part of the Bretagne BRE region and the western part of the Centre-Val de Loire CVL region. Low-risk areas (blue contours) were concentrated in the southwestern regions, including Occitanie and Nouvelle-Aquitaine (Figure 4A). GAM model predictions (Figure 4B) were consistent with the relative risk map, showing the highest *B. garinii* infection probabilities in the identified high-risk areas. Additional suitable areas for *B. garinii* were predicted in northwestern France, particularly in Bretagne BRE, Normandie NOR, and Hauts-de-France HDF, as well as in the northern parts of Nouvelle-Aquitaine NAQ and Centre-Val de Loire CVL. Uncertainty associated with the infection probability predictions was generally low but increased in areas with the highest predicted infection probability, notably in the high-risk zones highlighted on the map (Figure 4C).

## Discussion

Assessing spatial risk of Lyme borreliosis is challenging, as localized surveys lack coverage and broad incidence data miss ecological drivers. By leveraging CiTIQUE citizen-science data, our study provides the first geographically explicit, large-scale view of *Borrelia burgdorferi* sensu lato distribution in France, linking infection risk with key environmental, ecological, and anthropogenic factors.

### Spatial heterogeneity of the tick infection rates

*B. afzelii* and *B. garinii* were the most prevalent genospecies in the biting ticks which is consistent with the dominant genospecies and their relative frequencies reported across European countries, primarily based on questing tick studies (5, 79). However, the pathogen detection method only identified the dominant genospecies in co-infected ticks, potentially underestimating the prevalence of other species. For example, if *B. afzelii* dominates in co-infected ticks, it may have been disproportionately reported in areas with high tick abundance and higher frequencies of co-infection.

Previous work at the European scale (7), using a rough resolution (around 28 km × 28 km per cell) reported high prevalence of *B. afzelii* across France, particularly in eastern Auvergne-Rhône-Alpes. In contrast, our analyses at a finer resolution (5km x5km per cells) revealed a more heterogeneous pattern, with higher infection rates in eastern and central regions (Grand Est, Bourgogne-Franche-Comté, Centre-Val de Loire) and lower rates in western, northern, and southern regions (Bretagne, Normandie, Occitanie). For *B. garinii*, they found very high prevalence in these western, southern and northern regions, whereas our analyses revealed a more even distribution nationwide, while still confirming higher infection rates in the western and northern part.

### Influence of habitats and hosts on tick infection rates

Our GAMs highlighted distinct environmental and ecological factors associated with the distribution of Bbsl and its two main genospecies, *B. afzelii* and *B. garinii*. At the overall Bbsl level, infection probability was positively associated with the *I. ricinus* habitat suitability index, a composite indicator derived from a multi-criteria analysis integrating climate, altitude, land cover and wild ungulates density, which are factors known to influence tick abundance (21). This result suggests that, in areas where human exposure occurs, favourable environments for ticks also promote higher Bbsl circulation and prevalence. This finding aligns with theoretical models predicting a positive, yet non-linear, relationship between tick density and pathogen prevalence, modulated by host community composition, including the presence of non-competent host for Bbsl transmission (80, 81).

Observational studies in comparable ecological settings are consistent with these results. In Belgium, tick density was identified as a key driver of Bbsl-infected nymph density, with limited evidence for dilution effects from non-competent hosts (82). Similarly, in the Netherlands, increased ungulate abundance led to higher tick density and a non-linear, accelerating rise in the density of Bbsl-infected nymphs, without clear evidence for dilution effects attributable to ungulates (83). Long-term monitoring in southern England further showed that habitats favourable to ticks, such as structurally diverse forests and woodland edges, sustain higher densities of both questing and Bbsl-infected *I. ricinus* nymphs (84).

At the genospecies level, habitat and host associations differed, reflecting the distinct ecological characteristics of *B. afzelii* and *B. garinii* (6, 85, 86). *Borrelia garinii* infection probability was positively associated with Turdidae bird abundance, consistent with its known reliance on avian hosts (87, 88), and negatively associated with rodent species richness, possibly reflecting a dilution effect from these non-specific host (83). Conversely, *B. afzelii* infection probability was positively associated with rodent richness, consistent with the role of small mammals as primary reservoirs for this genospecies (39). Rodent richness was also positively associated with the presence of Bbsl, likely reflecting both the dominance of *B. afzelii* in our dataset and the association of several minor Bbsl genospecies, as *B. burgdorferi sensu stricto*, with small mammal hosts (89). However these results on *B. afzelii* could be either because *B. afzelii* is very dominant especially in the core areas of *Borrelia* presence or it could be a consequence of a method bias toward *B. afzelii* in co infected ticks, linking its presence with areas of high infection rate.

Grass cover exhibited a non-linear association with Bbsl and *B. afzelii* infection probabilities, with a negative trend across most observed values. Elevated risk was observed at low grass cover levels, characteristic of forest, fragmented woodlands, and associated ecotones, including adjacent private gardens, which are all known to favour both tick and *Borrelia* presence (90). In contrast, extensive grass cover, typical of open meadows or pastures, was associated with lower infection probabilities, likely due to reduced tick survival in open, hot, and dry environments, and a lower density of competent hosts (91, 92).

### Present limitations and perspectives offered by the continuous engagement of citizens

Our modelling approach prioritised interpretability using a parsimonious set of covariates, selected to limit collinearity and capture the main ecological drivers, while accounting for the current sampling structure. Although additional covariates, such as vegetation indices (7), forest structural complexity (93) or soil properties (94), have been highlighted in previous studies, our sampling resolution likely limits the reliable detection of such fine-scale environmental effects.

Sampling constraints also restricted our capacity to investigate several relevant aspects of Bbsl infection risk, including variation across tick developmental stages, seasonal and inter-annual dynamics, as well as infection rates across specific environmental contexts. Despite efforts to mitigate sampling bias, by analysing a minimum of 150 ticks per region, a substantial proportion of the analysed samples originated from densely populated areas, leaving gaps in remote and sparsely populated regions in France. A further challenge in estimating Lyme disease risk is the limited understanding of the components leading host seeking infected Bbsl tick to human populations (95). In this regard, our analysis on human-biting ticks provides valuable insights (96). However, a more precise assessment of risk would require comparison with infection rates in questing ticks, to disentangle ecological hazard from human exposure. Finally, integrating CiTIQUE tick-bite reports could provide complementary information on the socio-behavioral and ecological factors shaping human exposure thereby improving risk mapping (97).

Despite these limitations, our study demonstrates the substantial potential of citizen-generated data for monitoring tick-borne pathogen distribution at a national scale. The ticks analysed here represent approximately 40 % of all human-biting ticks submitted to CiTIQUE along with a tick-bite report during the study period. Since its inception, the CiTIQUE programme has grown considerably, with over 60,000 human and animal-biting ticks currently stored in the national tick bank. This continuously expanding resource offers unique opportunities to refine surveillance and research on tick-borne diseases in France.

As data and biting ticks continues to be collected, targeted sub-sampling strategies could be implemented to improve spatial representativeness, assess temporal dynamics, compare infection rates across tick developmental stages, and address refined ecological questions regarding Bbsl infection risk. Such approaches will contribute to a more detailed understanding of the eco-epidemiology of Bbsl genospecies and other tick-borne pathogens in human-biting ticks.

In the context of global environmental change and potential shifts in ticks and tick-borne pathogen distributions, the continued development of CiTIQUE provides a scalable, adaptable, citizen-driven tool to support large scale surveillance, environmental risk assessment, and public health preparedness.

## Acknowledgement

Our thoughts are with Jean-François Cosson and Béatrice Palin, who both passed away. Jean-François Cosson co-created the CiTIQUE project in 2016 with Pascale Frey-Klett and Muriel Vayssier-Taussat. Béatrice Palin was the first “tick librarian” of the program. We extend our gratitude to the current and former members of the CiTIQUE team, particularly Irene Carravieri, Cyril Galley, Julien Marchand, Sandrine Capizzi, Gwenaël Vourc’h, Philippe Lecomte, Sara Moutailler, Clémence Galon and all the tick librarians, as well as to all the citizens and CiTIQUE partners who made this study possible by reporting tick bites and sending the collected ticks to the tick library.

We are also grateful for the significant financial support that helped collecting ticks and providing tick analyses, including from INRAE, ANSES, the Ministry of Health and Access to Care, the Laboratory of Excellence ARBRE (ANR-11-LABX-0002-01), the Grand Est Region, the European Regional Development Fund, the “Des Hommes et Des Arbres” investment program (Territoire d’Innovation), the Groupama Foundation, and the Fondation de France.

The post-doctoral grant of Thierno Madiou Bah was supported by the INRAE scientific divisions Animal Health and Ecology and Biodiversity.

## Declaration of generative AI in scientific writing

During the preparation of this work the authors used ChatGPT in order to improve English writing. After using this tool/service, the authors reviewed and edited the content as needed and take full responsibility for the content of the published article.

## Data availability

The dataset will be made publicly available through the Recherche Data Gouv repository (https://entrepot.recherche.data.gouv.fr) upon publication of this study.

## Ethics approval

Approval of the ethics committee was not required since participation in sending ticks was voluntary and the short questionnaire online was anonymous (no identification possible). The participants are given information about the study on the website of the project.

## Notes

### Competing Interest Statement

The authors have declared no competing interest.

### Summary of Updates

The authors affiliations were updated, and an Importance section as well as a declaration of generative AI use in scientific writing were added. The Introduction and Discussion sections were slightly shortened while maintaining their flow and meaning.

## References

1. Burn L, Tran TMP, Pilz A, Vyse A, Fletcher MA, Angulo FJ, Gessner BD, Moïsi JC, Jodar L, Stark JH. 2023. Incidence of Lyme Borreliosis in Europe from National Surveillance Systems (2005– 2020). Vector Borne Zoonotic Dis 23:156.

2. Suhanda IE, Le Bel J, Bonnet C, Launay T, Kim Y, Delory T, Métras R. 2025. Computerised decision support system towards informing Lyme borreliosis incidence in France. Sci Rep 15:15506.

3. Rizzoli A, Hauffe HC, Carpi G, Vourc’h GI, Neteler M, Rosà R. 2011. Lyme borreliosis in Europe. Eurosurveillance 16:19906.

4. Coburn J, Garcia B, Hu LT, Jewett MW, Kraiczy P, Norris SJ, Skare J. 2021. Lyme Disease Pathogenesis. Curr Issues Mol Biol 42:473–518.

5. Rauter C, Hartung T. 2005. Prevalence of Borrelia burgdorferi Sensu Lato Genospecies in Ixodes ricinus Ticks in Europe: a Metaanalysis. Appl Environ Microbiol 71:7203–7216.

6. Wolcott KA, Margos G, Fingerle V, Becker NS. 2021. Host association of Borrelia burgdorferi sensu lato: A review. Ticks Tick-Borne Dis 12:101766.

7. Estrada-Peña A, Cutler S, Potkonjak A, Vassier-Tussaut M, Van Bortel W, Zeller H, Fernández-Ruiz N, Mihalca AD. 2018. An updated meta-analysis of the distribution and prevalence of Borrelia burgdorferi s.l. in ticks in Europe. Int J Health Geogr 17:41.

8. Stanek G, Wormser GP, Gray J, Strle F. 2012. Lyme borreliosis. The Lancet 379:461–473.

9. Eisen L, Eisen RJ. 2016. Critical Evaluation of the Linkage Between Tick-Based Risk Measures and the Occurrence of Lyme Disease Cases. J Med Entomol 53:1050–1062.

10. Tjaden NB, Caminade C, Beierkuhnlein C, Thomas SM. 2018. Mosquito-Borne Diseases: Advances in Modelling Climate-Change Impacts. Trends Parasitol 34:227–245.

11. Gaff H, Eisen RJ, Eisen L, Nadolny R, Bjork J, Monaghan AJ. 2020. LYMESIM 2.0: An Updated Simulation of Blacklegged Tick (Acari: Ixodidae) Population Dynamics and Enzootic Transmission of Borrelia burgdorferi (Spirochaetales: Spirochaetaceae). J Med Entomol 57:715–727.

12. Tran T, Prusinski MA, White JL, Falco RC, Kokas J, Vinci V, Gall WK, Tober KJ, Haight J, Oliver J, Sporn LA, Meehan L, Banker E, Backenson PB, Jensen ST, Brisson D. 2022. Predicting spatiotemporal population patterns of Borrelia burgdorferi, the Lyme disease pathogen. J Appl Ecol 59:2779–2789.

13. Estrada-Peña A, Sánchez N, Estrada-Sánchez A. 2012. An Assessment of the Distribution and Spread of the Tick Hyalomma marginatum in the Western Palearctic Under Different Climate Scenarios. Vector-Borne Zoonotic Dis 12:758–768.

14. Rataud A, Galon C, Bournez L, Henry P-Y, Marsot M, Moutailler S. 2022. Diversity of Tick-Borne Pathogens in Tick Larvae Feeding on Breeding Birds in France. 8. Pathogens 11:946.

15. Wongnak P, Bord S, Jacquot M, Agoulon A, Beugnet F, Bournez L, Cèbe N, Chevalier A, Cosson J-F, Dambrine N, Hoch T, Huard F, Korboulewsky N, Lebert I, Madouasse A, Mårell A, Moutailler S, Plantard O, Pollet T, Poux V, René-Martellet M, Vayssier-Taussat M, Verheyden H, Vourc’h G, Chalvet-Monfray K. 2022. Meteorological and climatic variables predict the phenology of Ixodes ricinus nymph activity in France, accounting for habitat heterogeneity. Sci Rep 12:7833.

16. Perez G, Bournez L, Boulanger N, Fite J, Livoreil B, McCoy KD, Quillery E, René-Martellet M, Bonnet SI. 2023. The distribution, phenology, host range and pathogen prevalence of Ixodes ricinus in France: a systematic map and narrative review. Peer Community J 3:e81.

17. Septfons A, Goronflot T, Jaulhac B, Roussel V, Martino SD, Guerreiro S, Launay T, Fournier L, Valk HD, Figoni J, Blanchon T, Couturier E. 2019. Epidemiology of Lyme borreliosis through two surveillance systems: the national Sentinelles GP network and the national hospital discharge database, France, 2005 to 2016. Eurosurveillance 24:1800134.

18. Eisen L, Eisen RJ. 2021. Benefits and Drawbacks of Citizen Science to Complement Traditional Data Gathering Approaches for Medically Important Hard Ticks (Acari: Ixodidae) in the United States. J Med Entomol 58:1–9.

19. Lernout T, De Regge N, Tersago K, Fonville M, Suin V, Sprong H. 2019. Prevalence of pathogens in ticks collected from humans through citizen science in Belgium. Parasit Vectors 12:550.

20. Joly D, Brossard T, Cardot H, Cavailhes J, Hilal M, Wavresky P. 2010. Les types de climats en France, une construction spatiale. Cybergeo Eur J Geogr 10.4000/cybergeo.23155.

21. Lebert I, Bord S, Saint-Andrieux C, Cassar E, Gasqui P, Beugnet F, Chalvet-Monfray K, Vanwambeke SO, Vourc’h G, René-Martellet M. 2022. Habitat suitability map of Ixodes ricinus tick in France using multi-criteria analysis. 1. Geospatial Health 17.

22. Ferquel E, Garnier M, Marie J, Bernède-Bauduin C, Baranton G, Pérez-Eid C, Postic D. 2006. Prevalence of Borrelia burgdorferi Sensu Lato and Anaplasmataceae Members in Ixodes ricinus Ticks in Alsace, a Focus of Lyme Borreliosis Endemicity in France. Appl Environ Microbiol 72:3074–3078.

23. Vourc’h G, Abrial D, Bord S, Jacquot M, Masséglia S, Poux V, Pisanu B, Bailly X, Chapuis J-L. 2016. Mapping human risk of infection with Borrelia burgdorferi sensu lato, the agent of Lyme borreliosis, in a periurban forest in France. Ticks Tick-Borne Dis 7:644–652.

24. Nuttens C, Bessou A, Duret S, Skufca J, Blanc E, Pilz A, Gessner BD, Faucher J-F, Stark JH. 2023. Epidemiology of Lyme Borreliosis in France in Primary Care and Hospital Settings, 2010–2019. Vector-Borne Zoonotic Dis 23:221–229.

25. Durand J, Bah T-M, Lebert I, Galon C, Carravieri I, Masseglia S, Armand J-M, Marchand J, Galley C, Chalvet-Monfray K, Vayssier-Taussat M, Vour’ch G, Brun-Jacob A, Moutailler S, Bailly X, Frey-Klett P. 2025. Distribution of tick-borne pathogens in human-biting ticks in France collected through a Citizen-science program. medRxiv 10.1101/2025.09.22.25336155.

26. Pérez-Eid C. 2007. Les tiques1: identification, biologie, importance médicale et vétérinaire. Lavoisier.

27. Estrada-Peña A, Mihalca AD, Petney TN. 2018. Ticks of Europe and North Africa: A Guide to Species Identification. Springer.

28. Melis S, Batisti Biffignandi G, Olivieri E, Galon C, Vicari N, Prati P, Moutailler S, Sassera D, Castelli M. 2024. High-throughput screening of pathogens in Ixodes ricinus removed from hosts in Lombardy, northern Italy. Ticks Tick-Borne Dis 15:102285.

29. Michelet L, Delannoy S, Devillers E, Umhang G, Aspan A, Juremalm M, Chirico J, van der Wal FJ, Sprong H, Boye Pihl TP, Klitgaard K, Bødker R, Fach P, Moutailler S. 2014. High-throughput screening of tick-borne pathogens in Europe. Front Cell Infect Microbiol 4.

30. Bennet L, Halling A, Berglund J. 2006. Increased incidence of Lyme borreliosis in southern Sweden following mild winters and during warm, humid summers. Eur J Clin Microbiol Infect Dis 25:426–432.

31. Gilbert L, Maffey GL, Ramsay SL, Hester AJ. 2012. The effect of deer management on the abundance of in Scotland. Ecol Appl 22:658–667.

32. Fox J, Weisberg S. 2018. An R Companion to Applied Regression. SAGE Publications.

33. Mysterud A, Easterday WR, Stigum VM, Aas AB, Meisingset EL, Viljugrein H. 2016. Contrasting emergence of Lyme disease across ecosystems. Nat Commun 7:11882.

34. Król N, Obiegala A, Imholt C, Arz C, Schmidt E, Jeske K, Ulrich RG, RenteríalZJSolís Z, Jacob J, Pfeffer M. 2022. Diversity of Borrelia burgdorferi sensu lato in ticks and small mammals from different habitats. Parasit Vectors 15:195.

35. Ratti V, Winter JM, Wallace DI. 2021. Dilution and amplification effects in Lyme disease: Modeling the effects of reservoir-incompetent hosts on Borrelia burgdorferi sensu stricto transmission. Ticks Tick-Borne Dis 12:101724.

36. Jenkins CN, Pimm SL, Joppa LN. 2013. Global patterns of terrestrial vertebrate diversity and conservation. Proc Natl Acad Sci 110:E2602–E2610.

37. Sprong H, Moonen S, van Wieren SE, Hofmeester TR. 2020. Effects of cattle grazing on Ixodes ricinus-borne disease risk in forest areas of the Netherlands. Ticks Tick-Borne Dis 11:101355.

38. Robinson TP, Wint GRW, Conchedda G, Boeckel TPV, Ercoli V, Palamara E, Cinardi G, D’Aietti L, Hay SI, Gilbert M. 2014. Mapping the Global Distribution of Livestock. PLOS ONE 9:e96084.

39. Hanincová K, Schäfer SM, Etti S, Sewell H-S, Taragelová V, Ziak D, Labuda M, Kurtenbach K. 2003. Association of Borrelia afzelii with rodents in Europe. Parasitology 126:11–20.

40. Wint W, Morley D, Alexander N. 2013. Four rodent and vole biodiversity models for Europe. Open Heal Data 1:e3.

41. Gassner F, Takken W, Plas CL der, Kastelein P, Hoetmer AJ, Holdinga M, van Overbeek LS. 2013. Rodent species as natural reservoirs of Borrelia burgdorferi sensu lato in different habitats of Ixodes ricinus in The Netherlands. Ticks Tick-Borne Dis 4:452–458.

42. Dubska L, Literak I, Kocianova E, Taragelova V, Sychra O. 2009. Differential Role of Passerine Birds in Distribution of Borrelia Spirochetes, Based on Data from Ticks Collected from Birds during the Postbreeding Migration Period in Central Europe. Appl Environ Microbiol 75:596–602.

43. Pichon B, Rogers M, Egan D, Gray J. 2005. Blood-Meal Analysis for the Identification of Reservoir Hosts of Tick-Borne Pathogens in Ireland. Vector-Borne Zoonotic Dis 5:172–180.

44. Herrmann C, Gern L. 2010. Survival of Ixodes ricinus (Acari: Ixodidae) Under Challenging Conditions of Temperature and Humidity Is Influenced by Borrelia burgdorferi sensu lato Infection. J Med Entomol 47:1196–1204.

45. Shih CM, Telford SR, Spielman A. 1995. Effect of ambient temperature on competence of deer ticks as hosts for Lyme disease spirochetes. J Clin Microbiol 33:958–961.

46. Fick SE, Hijmans RJ. 2017. WorldClim 2: new 1-km spatial resolution climate surfaces for global land areas. Int J Climatol 37:4302–4315.

47. Ogden NH, Bigras-Poulin M, O’Callaghan CJ, Barker IK, Kurtenbach K, Lindsay LR, Charron DF. 2007. Vector seasonality, host infection dynamics and fitness of pathogens transmitted by the tick Ixodes scapularis. Parasitology 134:209–227.

48. Gatewood AG, Liebman KA, Vourc’h G, Bunikis J, Hamer SA, Cortinas R, Melton F, Cislo P, Kitron U, Tsao J, Barbour AG, Fish D, Diuk-Wasser MA. 2009. Climate and Tick Seasonality Are Predictors of Borrelia burgdorferi Genotype Distribution. Appl Environ Microbiol 75:2476–2483.

49. Kiewra D, Kryza M, Szymanowski M. 2014. Influence of selected meteorological variables on the questing activity of Ixodes ricinus ticks in Lower Silesia, SW Poland. J Vector Ecol 39:138– 145.

50. Hauser G, Rais O, Morán Cadenas F, Gonseth Y, Bouzelboudjen M, Gern L. 2018. Influence of climatic factors on Ixodes ricinus nymph abundance and phenology over a long-term monthly observation in Switzerland (2000–2014). Parasit Vectors 11:289.

51. Milne A. 1950. The ecology of the sheep tick, Ixodes ricinus L.: Microhabitat economy of the adult tick. Parasitology 40:14–34.

52. Berger KA, Ginsberg HS, Dugas KD, Hamel LH, Mather TN. 2014. Adverse moisture events predict seasonal abundance of Lyme disease vector ticks (Ixodes scapularis). Parasit Vectors 7:181.

53. Burtis JC, Sullivan P, Levi T, Oggenfuss K, Fahey TJ, Ostfeld RS. 2016. The impact of temperature and precipitation on blacklegged tick activity and Lyme disease incidence in endemic and emerging regions. Parasit Vectors 9:606.

54. Verhulst J, Báldi A, Kleijn D. 2004. Relationship between land-use intensity and species richness and abundance of birds in Hungary. Agric Ecosyst Environ 104:465–473.

55. Nikolov SC, Demerdzhiev DA, Popgeorgiev GS, Plachiyski DG. 2011. Bird community patterns in sub–Mediterranean pastures: the effects of shrub cover and grazing intensity. 1. Anim Biodivers Conserv 34:11–21.

56. Thompson SJ, Handel CM, Richardson RM, McNew LB. 2016. When Winners Become Losers: Predicted Nonlinear Responses of Arctic Birds to Increasing Woody Vegetation. PLOS ONE 11:e0164755.

57. Buchhorn M, Smets B, Bertels L, Roo BD, Lesiv M, Tsendbazar N-E, Herold M, Fritz S. 2020. Copernicus Global Land Service: Land Cover 100m: collection 3: epoch 2018: Globe (V3.0.1). Zenodo.

58. Tack W, Madder M, Baeten L, De Frenne P, Verheyen K. 2012. The abundance of Ixodes ricinus ticks depends on tree species composition and shrub cover. Parasitology 139:1273–1281.

59. Ruyts SC, Tack W, Ampoorter E, Coipan EC, Matthysen E, Heylen D, Sprong H, Verheyen K. 2018. Year-to-year variation in the density of Ixodes ricinus ticks and the prevalence of the rodent-associated human pathogens Borrelia afzelii and B. miyamotoi in different forest types. Ticks Tick-Borne Dis 9:141–145.

60. Perez G, Bastian S, Chastagner A, Agoulon A, Rantier Y, Vourc’h G, Plantard O, Butet A. 2020. Relationships between landscape structure and the prevalence of two tick-borne infectious agents, Anaplasma phagocytophilum and Borrelia burgdorferi sensu lato, in small mammal communities. Landsc Ecol 35:435–451.

61. Oechslin CP, Heutschi D, Lenz N, Tischhauser W, Péter O, Rais O, Beuret CM, Leib SL, Bankoul S, Ackermann-Gäumann R. 2017. Prevalence of tick-borne pathogens in questing Ixodes ricinus ticks in urban and suburban areas of Switzerland. Parasit Vectors 10:558.

62. Heylen D, Lasters R, Adriaensen F, Fonville M, Sprong H, Matthysen E. 2019. Ticks and tickborne diseases in the city: Role of landscape connectivity and green space characteristics in a metropolitan area. Sci Total Environ 670:941–949.

63. Mathews-Martin L, Namèche M, Vourc’h G, Gasser S, Lebert I, Poux V, Barry S, Bord S, Jachacz J, Chalvet-Monfray K. 2020. Questing tick abundance in urban and peri-urban parks in the French city of Lyon. Parasit Vectors 13:1–9.

64. Hansford KM, Gillingham EL, Vaux AGC, Cull B, McGinley L, Catton M, Wheeler BW, Tschirren B, Medlock JM. 2023. Impact of green space connectivity on urban tick presence, density and Borrelia infected ticks in different habitats and seasons in three cities in southern England. Ticks Tick-Borne Dis 14:102103.

65. Weiss D. 2024. A global map of travel time to cities. DANS Data Station Physical and Technical Sciences.

66. Skinner EB, Glidden CK, MacDonald AJ, Mordecai EA. 2023. Human footprint is associated with shifts in the assemblages of major vector-borne diseases. Nat Sustain 6:652–661.

67. Mu H, Li X, Wen Y, Huang J, Du P, Su W, Miao S, Geng M. 2022. A global record of annual terrestrial Human Footprint dataset from 2000 to 2018. Sci Data 9:176.

68. Wardle DA, Bardgett RD, Klironomos JN, Setälä H, van der Putten WH, Wall DH. 2004. Ecological Linkages Between Aboveground and Belowground Biota. Science 304:1629–1633.

69. Lavelle P, Decaëns T, Aubert M, Barot S, Blouin M, Bureau F, Margerie P, Mora P, Rossi J-P. 2006. Soil invertebrates and ecosystem services. Eur J Soil Biol 42:S3–S15.

70. Burtis JC, Yavitt JB, Fahey TJ, Ostfeld RS. 2019. Ticks as Soil-Dwelling Arthropods: An Intersection Between Disease and Soil Ecology. J Med Entomol 56:1555–1564.

71. Orgiazzi A, Bardgett RD, Barrios E. 2016. Global soil biodiversity atlas.

72. Marra G, Wood SN. 2011. Practical variable selection for generalized additive models. Comput Stat Data Anal 55:2372–2387.

73. Wood SN. 2011. Fast Stable Restricted Maximum Likelihood and Marginal Likelihood Estimation of Semiparametric Generalized Linear Models. J R Stat Soc Ser B Stat Methodol 73:3–36.

74. Hartig F, Lohse L, leite M de S. 2024. DHARMa: Residual Diagnostics for Hierarchical (MultiLevel / Mixed) Regression Models (0.4.7).

75. Davies TM, Marshall JC, Hazelton ML. 2018. Tutorial on kernel estimation of continuous spatial and spatiotemporal relative risk. Stat Med 37:1191–1221.

76. Davies TM, Hazelton ML. 2010. Adaptive kernel estimation of spatial relative risk. Stat Med 29:2423–2437.

77. Diggle P. 1985. A Kernel Method for Smoothing Point Process Data. J R Stat Soc Ser C Appl Stat 34:138–147.

78. R Core Team. 2023. R: A Language and Environment for Statistical Computing. R Foundation for Statistical Computing, Vienna, Austria. https://www.R-project.org/.

79. Strnad M, Hönig V, Růžek D, Grubhoffer L, Rego ROM. 2017. Europe-Wide Meta-Analysis of Borrelia burgdorferi Sensu Lato Prevalence in Questing Ixodes ricinus Ticks. Appl Environ Microbiol 83:e00609–17.

80. Hartemink NA, Randolph SE, Davis SA, Heesterbeek J a. P. 2008. The Basic Reproduction Number for Complex Disease Systems: Defining R0 for Tick-Borne Infections. Am Nat 171:743–754.

81. Dagostin F, Tagliapietra V, Marini G, Cataldo C, Bellenghi M, Pizzarelli S, Cammarano RR, Wint W, Alexander NS, Neteler M, Haas J, Dub T, Busani L, Rizzoli A. 2023. Ecological and environmental factors affecting the risk of tick-borne encephalitis in Europe, 2017 to 2021. Eurosurveillance 28:2300121.

82. Ruyts SC, Landuyt D, Ampoorter E, Heylen D, Ehrmann S, Coipan EC, Matthysen E, Sprong H, Verheyen K. 2018. Low probability of a dilution effect for Lyme borreliosis in Belgian forests. Ticks Tick-Borne Dis 9:1143–1152.

83. Takumi K, Sprong H, Hofmeester TR. 2019. Impact of vertebrate communities on Ixodes ricinus-borne disease risk in forest areas. Parasit Vectors 12:434.

84. Medlock JM, Vaux AGC, Gandy S, Cull B, McGinley L, Gillingham E, Catton M, Pullan ST, Hansford KM. 2022. Spatial and temporal heterogeneity of the density of Borrelia burgdorferi-infected Ixodes ricinus ticks across a landscape: A 5-year study in southern England. Med Vet Entomol 36:356–370.

85. Gern L. 2008. Borrelia burgdorferi sensu lato, the agent of Lyme borreliosis: life in the wilds. 3. Parasite 15:244–247.

86. Fabri ND, Heesterbeek H, Cromsigt JPGM, Ecke F, Sprong H, Nijhuis L, Hofmeester TR, Hartemink N. 2024. Exploring the influence of host community composition on the outbreak po-tential of Anaplasma phagocytophilum and Borrelia burgdorferi s.l. Ticks Tick-Borne Dis 15:102275.

87. Norte AC, Margos G, Becker NS, Albino Ramos J, Núncio MS, Fingerle V, Araújo PM, Adamík P, Alivizatos H, Barba E, Barrientos R, Cauchard L, Csörgő T, Diakou A, Dingemanse NJ, Doligez B, Dubiec A, Eeva T, Flaisz B, Grim T, Hau M, Heylen D, Hornok S, Kazantzidis S, Kováts D, Krause F, Literak I, Mänd R, Mentesana L, Morinay J, Mutanen M, Neto JM, Nováková M, Sanz JJ, Pascoal da Silva L, Sprong H, Tirri I-S, Török J, Trilar T, Tyller Z, Visser ME, Lopes de Carvalho I. 2020. Host dispersal shapes the population structure of a tick-borne bacterial pathogen. Mol Ecol 29:485–501.

88. Taragel’ová V, Koči J, Hanincová K, Kurtenbach K, Derdáková M, Ogden NH, Literák I, Kocianová E, Labuda M. 2008. Blackbirds and Song Thrushes Constitute a Key Reservoir of Borrelia garinii, the Causative Agent of Borreliosis in Central Europe. Appl Environ Microbiol 74:1289–1293.

89. Margos G, Vollmer SA, Ogden NH, Fish D. 2011. Population genetics, taxonomy, phylogeny and evolution of Borrelia burgdorferi sensu lato. Infect Genet Evol 11:1545–1563.

90. Bourdin A, Dokhelar T, Bord S, van Halder I, Stemmelen A, Scherer-Lorenzen M, Jactel H. 2023. Forests harbor more ticks than other habitats: A meta-analysis. For Ecol Manag 541:121081.

91. Guerra M, Walker E, Jones C, Paskewitz S, Cortinas MR, Stancil A, Beck L, Bobo M, Kitron U. 2002. Predicting the Risk of Lyme Disease: Habitat Suitability for Ixodes scapularis in the North Central United States. Emerg Infect Dis 8:289–297.

92. Boyard C, Vourc’h G, Barnouin J. 2008. The relationships between Ixodes ricinus and small mammal species at the woodland–pasture interface. Exp Appl Acarol 44:61–76.

93. Vanroy T, Martel A, Baeten L, Fonville M, Lens L, Pasmans F, Sprong H, Strubbe D, Van Gestel M, Verheyen K. 2024. The effect of forest structural complexity on tick-borne pathogens in questing ticks and small mammals. For Ecol Manag 562:121944.

94. Burtis JC, Yavitt JB, Fahey TJ, Ostfeld RS. 2019. Ticks as Soil-Dwelling Arthropods: An Intersection Between Disease and Soil Ecology. J Med Entomol 56:1555–1564.

95. Eisen L, Eisen RJ. 2016. Critical Evaluation of the Linkage Between Tick-Based Risk Measures and the Occurrence of Lyme Disease Cases. J Med Entomol 53:1050–1062.

96. Hook SA, Nawrocki CC, Meek JI, Feldman KA, White JL, Connally NP, Hinckley AF. 2021. Human-tick encounters as a measure of tickborne disease risk in lyme disease endemic areas. Zoonoses Public Health 68:384–392.

97. Bouchard C, Dumas A, Baron G, Bowser N, Leighton PA, Lindsay LR, Milord F, Ogden NH, Aenishaenslin C. 2023. Integrated human behavior and tick risk maps to prioritize Lyme disease interventions using a “One Health” approach. Ticks Tick-Borne Dis 14:102083.

