## Supplemental Table 1 for "Infection rate of *Borrelia burgdorferi* genospecies in human-biting *Ixodes ricinus* ticks: models for surveillance based on the French citizen science programme CiTIQUE"

Supplementary Table 1 Covariates extracted for the analysis with their sources and resolution.

| Type | name | Variable | Source | Resolution |
| --- | --- | --- | --- | --- |
| Bioclimatic | copernicus_DSM_30arcsec_mood_bbox_epsg4326 | Altitude | Copernicus Digital Elevation Model | 30s |
| Bioclimatic | wc2.1_30s_bio_1 | Annual Mean Temperature | Worldcilm 2 (Fick and Hijmans, 2017) | 30s |
| Bioclimatic | wc2.1_30s_bio_2 | Mean Diurnal Range (Mean of monthly (max temp - min temp)) | Worldcilm 2 (Fick and Hijmans, 2017) | 30s |
| Bioclimatic | wc2.1_30s_bio_3 | Isothermality (BIO2/BIO7) (×100) | Worldcilm 2 (Fick and Hijmans, 2017) | 30s |
| Bioclimatic | wc2.1_30s_bio_4 | Temperature Seasonality (standard deviation ×100) | Worldcilm 2 (Fick and Hijmans, 2017) | 30s |
| Bioclimatic | wc2.1_30s_bio_5 | Max Temperature of Warmest Month | Worldcilm 2 (Fick and Hijmans, 2017) | 30s |
| Bioclimatic | wc2.1_30s_bio_6 | Min Temperature of Coldest Month | Worldcilm 2 (Fick and Hijmans, 2017) | 30s |
| Bioclimatic | wc2.1_30s_bio_7 | Temperature Annual Range (BIO5-BIO6) | Worldcilm 2 (Fick and Hijmans, 2017) | 30s |
| Bioclimatic | wc2.1_30s_bio_8 | Mean Temperature of Wettest Quarter | Worldcilm 2 (Fick and Hijmans, 2017) | 30s |
| Bioclimatic | wc2.1_30s_bio_9 | Mean Temperature of Driest Quarter | Worldcilm 2 (Fick and Hijmans, 2017) | 30s |
| Bioclimatic | wc2.1_30s_bio_10 | Mean Temperature of Warmest Quarter | Worldcilm 2 (Fick and Hijmans, 2017) | 30s |
| Bioclimatic | wc2.1_30s_bio_11 | Mean Temperature of Coldest Quarter | Worldcilm 2 (Fick and Hijmans, 2017) | 30s |
| Bioclimatic | wc2.1_30s_bio_12 | Annual Precipitation | Worldcilm 2 (Fick and Hijmans, 2017) | 30s |
| Bioclimatic | wc2.1_30s_bio_13 | Precipitation of Wettest Month | Worldcilm 2 (Fick and Hijmans, 2017) | 30s |
| Bioclimatic | wc2.1_30s_bio_14 | Precipitation of Driest Month | Worldcilm 2 (Fick and Hijmans, 2017) | 30s |
| Bioclimatic | wc2.1_30s_bio_15 | Precipitation Seasonality (Coefficient of Variation) | Worldcilm 2 (Fick and Hijmans, 2017) | 30s |
| Bioclimatic | wc2.1_30s_bio_16 | Precipitation of Wettest Quarter | Worldcilm 2 (Fick and Hijmans, 2017) | 30s |
| Bioclimatic | wc2.1_30s_bio_17 | Precipitation of Driest Quarter | Worldcilm 2 (Fick and Hijmans, 2017) | 30s |
| Bioclimatic | wc2.1_30s_bio_18 | Precipitation of Warmest Quarter | Worldcilm 2 (Fick and Hijmans, 2017) | 30s |
| Bioclimatic | wc2.1_30s_bio_19 | Precipitation of Coldest Quarter | Worldcilm 2 (Fick and Hijmans, 2017) | 30s |
| Bioclimatic | Daylength | Daylength | geosphere (Forsythe et al., 1995) |  |
| Bioclimatic | Day of the year | Day of the year |  |  |
| Bioclimatic | cos day of the year | Cosinus of day of the year cos(2*pi*day_of_the_year/365) |  |  |
| Bioclimatic | sin day of the year | Sinus of day of the year sin(2*pi*day_of_the_year/365) |  |  |
| Landcover | PROBAV_LC100_global_v3.0.1_2019_nrt_Bare_CoverFraction_layer_EPSG_4326 | Bare_Cover_Fraction | Copernicus Global Land Service (Buchhorn et al., 2020) | 100m |
| Landcover | PROBAV_LC100_global_v3.0.1_2019_nrt_BuiltUp_CoverFraction_layer_EPSG_4326 | Built_Up_Cover_Fraction | Copernicus Global Land Service (Buchhorn et al., 2020) | 100m |
| Landcover | PROBAV_LC100_global_v3.0.1_2019_nrt_Crops_CoverFraction_layer_EPSG_4326 | Crops_Cover_Fraction | Copernicus Global Land Service (Buchhorn et al., 2020) | 100m |
| Landcover | PROBAV_LC100_global_v3.0.1_2019_nrt_Grass_CoverFraction_layer_EPSG_4326 | Grass_Cover_Fraction | Copernicus Global Land Service (Buchhorn et al., 2020) | 100m |
| Landcover | PROBAV_LC100_global_v3.0.1_2019_nrt_PermanentWater_CoverFraction_layer_EPSG_4326 | Permanent_Water_Cover_Fraction | Copernicus Global Land Service (Buchhorn et al., 2020) | 100m |
| Landcover | PROBAV_LC100_global_v3.0.1_2019_nrt_SeasonalWater_CoverFraction_layer_EPSG_4326 | Seasonal_Water_Cover_Fraction | Copernicus Global Land Service (Buchhorn et al., 2020) | 100m |
| Landcover | PROBAV_LC100_global_v3.0.1_2019_nrt_Shrub_CoverFraction_layer_EPSG_4326 | Shrub_Cover_Fraction | Copernicus Global Land Service (Buchhorn et al., 2020) | 100m |
| Landcover | PROBAV_LC100_global_v3.0.1_2019_nrt_Tree_CoverFraction_layer_EPSG_4326 | Tree_Cover_Fraction | Copernicus Global Land Service (Buchhorn et al., 2020) | 100m |
| Landcover | PROBAV_LC100_global_v3.0.1_2019_nrt_Forest_Type_layer_EPSG_4326 | Forest type (ENF, EBF, DNF ,DBF, Mixed) for each Tree cover Fraction that exceed 1% | Copernicus Global Land Service (Buchhorn et al., 2020) | 100m |
| Landcover | Sand1 | Topsoil sand composition | Topsoil physical properties for Europe ESDAC (Ballabio et al., 2016) | 500m |
| Landcover | Silt1 | Topsoil silt composition | Topsoil physical properties for Europe ESDAC (Ballabio et al., 2016) | 500m |
| Landcover | mo1115a0cog | Fourier Processed MODIS EVI Enhanced_Vegetation Index_2001 2021_mean | MODIS (Scharlemann et al., 2008) | 1km |
| Landcover | mo1115a1cog | Fourier Processed MODIS EVI Enhanced_Vegetation Index_2001 2021_amplitude_1 | MODIS (Scharlemann et al., 2008) | 1km |
| Landcover | mo1115a2cog | Fourier Processed MODIS EVI Enhanced_Vegetation Index_2001 2021_amplitude_2 | MODIS (Scharlemann et al., 2008) | 1km |
| Landcover | mo1115a3cog | Fourier Processed MODIS EVI Enhanced_Vegetation Index_2001 2021_amplitude_3 | MODIS (Scharlemann et al., 2008) | 1km |
| Landcover | mo1115mncog | Fourier Processed MODIS EVI Enhanced_Vegetation Index_2001 2021_min | MODIS (Scharlemann et al., 2008) | 1km |
| Landcover | mo1115mxcog | Fourier Processed MODIS EVI Enhanced_Vegetation Index_2001 2021_max | MODIS (Scharlemann et al., 2008) | 1km |
| Landcover | mo1115p1cog | Fourier Processed MODIS EVI Enhanced_Vegetation Index_2001 2021_phase_1 | MODIS (Scharlemann et al., 2008) | 1km |
| Landcover | mo1115p2cog | Fourier Processed MODIS EVI Enhanced_Vegetation Index_2001 2021_phase_2 | MODIS (Scharlemann et al., 2008) | 1km |
| Landcover | mo1115p3cog | Fourier Processed MODIS EVI Enhanced_Vegetation Index_2001 2021_phase_3 | MODIS (Scharlemann et al., 2008) | 1km |
| Landcover | mo1115vrcog | Fourier Processed MODIS EVI Enhanced_Vegetation Index_2001 2021_variance | MODIS (Scharlemann et al., 2008) | 1km |
| Landcover | mo1114a0cog | Fourier Processed MODIS NDVI Enhanced_Vegetation Index_2001 2021_mean | MODIS (Scharlemann et al., 2008) | 1km |
| Landcover | mo1114a1cog | Fourier Processed MODIS NDVI Enhanced_Vegetation Index_2001 2021_amplitude_1 | MODIS (Scharlemann et al., 2008) | 1km |
| Landcover | mo1114a2cog | Fourier Processed MODIS NDVI Enhanced_Vegetation Index_2001 2021_amplitude_2 | MODIS (Scharlemann et al., 2008) | 1km |
| Landcover | mo1114a3cog | Fourier Processed MODIS NDVI Enhanced_Vegetation Index_2001 2021_amplitude_3 | MODIS (Scharlemann et al., 2008) | 1km |
| Landcover | mo1114mncog | Fourier Processed MODIS NDVI Enhanced_Vegetation Index_2001 2021_min | MODIS (Scharlemann et al., 2008) | 1km |
| Landcover | mo1114mxcog | Fourier Processed MODIS NDVI Enhanced_Vegetation Index_2001 2021_max | MODIS (Scharlemann et al., 2008) | 1km |
| Landcover | mo1114p1cog | Fourier Processed MODIS NDVI Enhanced_Vegetation Index_2001 2021_phase_1 | MODIS (Scharlemann et al., 2008) | 1km |
| Landcover | mo1114p2cog | Fourier Processed MODIS NDVI Enhanced_Vegetation Index_2001 2021_phase_2 | MODIS (Scharlemann et al., 2008) | 1km |
| Landcover | mo1114p3cog | Fourier Processed MODIS NDVI Enhanced_Vegetation Index_2001 2021_phase_3 | MODIS (Scharlemann et al., 2008) | 1km |
| Landcover | mo1114vrcog | Fourier Processed MODIS NDVI Enhanced_Vegetation Index_2001 2021_variance | MODIS (Scharlemann et al., 2008) | 1km |
| Human pressure | eraccess505km | Accessibility to high-density urban centres in 2015 | (Weiss, 2024) | 1km |
| Human pressure | hfp2019 | 2019 global human footprint | (Mu et al., 2022) | 1km |
| Human pressure | hfp2020 | 2020 global human footprint | (Mu et al., 2022) | 1km |
| Human pressure | vcwpopppp | Human population density | Worldpop | 1km |
| Human pressure | soil_bio_new | Soil biodiversity | ESDAC (Orgiazzi et al., 2016) | 10km |
| Human pressure | threat_soil_bio | Soil biodiversity threats | ESDAC (Orgiazzi et al., 2016) | 10km |
| Human pressure | MONODWATALLVETSNODUPMAY21 | Healthcare and veterinary locations | ERGO MOOD project William Wint (undisclosed data) |  |
| Vectors | Ad418Dpt_R100 | Habitat suitability index for *I. ricinus* | (Lebert et al., 2022) | 100m |
| Vectors | ixricnewsuitensrfbrt15kbaldec19 | Proportion of suitable habitat for *I. ricinus* | ERGO MOOD project William Wint (undisclosed data) | 1km |
| Hosts | all3deeravprob | Sum of proportion suitable habitat for roe deer, red deer and fallow deer | ERGO MOOD project William Wint (undisclosed data) | 1km |
| Hosts | ensredmodel | Proportion of suitable habitat for red deer | ERGO MOOD project William Wint (undisclosed data) | 1km |
| Hosts | ensroemodel | Proportion of suitable habitat for roe deer | ERGO MOOD project William Wint (undisclosed data) | 1km |
| Hosts | fallow_deer_damaRF_pa2_RFprobLW | Proportion of suitable habitat for fallow deer | ERGO MOOD project William Wint (undisclosed data) | 1km |
| Hosts | Anthus_trivialis_abundance | Abundance of *Anthus trivialis* | E-birds | 3km |
| Hosts | Cyanistes_caeruleus_abundance | Abundance *of Cyanistes caeruleus* | E-birds | 3km |
| Hosts | Erithacus_rubecula_abundance | Abundance of *Erithacus rubecula* | E-birds | 3km |
| Hosts | Ficedula_albicollis_abundance | Abundance of *Ficedula albicollis* | E-birds | 3km |
| Hosts | Luscinia_megarhynchos_abundance | Abundance of *Luscinia megarhynchos* | E-birds | 3km |
| Hosts | Parus_major_abundance | Abundance of *Parus major* | E-birds | 3km |
| Hosts | Phylloscopus_trochilus_abundance | Abundance of *Phylloscopus trochilus* | E-birds | 3km |
| Hosts | Prunella_modularis_abundance | Abundance of *Prunella modularis* | E-birds | 3km |
| Hosts | Sylvia_atricapilla_abundance | Abundance of *Sylvia atricapilla* | E-birds | 3km |
| Hosts | Troglodytes_troglodytes_abundance | Abundance of *Troglodytes troglodytes* | E-birds | 3km |
| Hosts | Turdus_iliacus_abundance | Abundance of *Turdus iliacus* | E-birds | 3km |
| Hosts | Turdus_merula_abundance | Abundance of *Turdus merula* | E-birds | 3km |
| Hosts | Turdus_philomelos_abundance | Abundance of *Turdus philomelos* | E-birds | 3km |
| Hosts | Turdus_pilaris_abundance | Abundance of *Turdus pilaris* | E-birds | 3km |
| Hosts | Cattle_2015_Da | Density of cattle per km² | Gridded livestock of the world database (Robinson et al., 2014) | 10km |
| Hosts | Goats_2015_Da | Density of goats per km² | Gridded livestock of the world database (Robinson et al., 2014) | 10km |
| Hosts | Horses_2015_Da | Density of horses per km² | Gridded livestock of the world database (Robinson et al., 2014) | 10km |
| Hosts | Sheep_2015_Da | Density of sheep per km² | Gridded livestock of the world database (Robinson et al., 2014) | 10km |
| Hosts | ergoapodemusfavicollisv2NLDA_AvProb | Proportion of suitable habitat for *apodemus favicollis* | ERGO MOOD project William Wint (undisclosed data) | 1km |
| Hosts | ergomyodesglareolus2NLDA_AvProb | Proportion of suitable habitat for *myodes glareolus* | ERGO MOOD project William Wint (undisclosed data) | 1km |
| Hosts | hant10 | Predicted average number of rodent species in the field between : *Apodemus agrarius*, *Apodemus flavicollis*, *Apodemus myst*, *Apodemus sylvatica*, *Myodes glariolus*, *Micotus arvensis*, *Microyus sylvatica*, *Rattus norwegica*, *Rattus rattus*, *Sorex araneus* and *Sorex minutus* | (Wint et al., 2013) | 1km |
| Hosts | sp04 | Predicted average number of rodent species in the field between : *Apodemus agrarius*, *Apodemus flavicollis*, *Myodes glareolus*, *Microtus arvensis* | (Wint et al., 2013) | 1km |
| Hosts | sp05 | *Predicted average number of rodent species in the field between : Apodemus agrarius, Apodemus flavicollis, Myodes glareolus, Microtus arvensis, Apodemus sylvaticus* | (Wint et al., 2013) | 1km |
| Hosts | ters04 | Predicted average number of rodent species in the field between : *Apodemus flavicollis*, *Microtus arvensis*, *Apodemus sylvaticus*, *Sorex araneus* | (Wint et al., 2013) | 1km |
| Hosts | Richness_10km_Birds_v7_EckertIV_Passeriformes | Passeriformes species richness | Biodiversity mapping (Jenkins et al., 2013) | 10km |
| Hosts | Richness_10km_MAMMALS_mar2018_EckertIV_Carnivora. | Carnivora species richness | Biodiversity mapping (Jenkins et al., 2013) | 10km |
| Hosts | Richness_10km_MAMMALS_mar2018_EckertIV_Cetartiodactyla | Cetartiodactyla species richness | Biodiversity mapping (Jenkins et al., 2013) | 10km |
| Hosts | Richness_10km_MAMMALS_mar2018_EckertIV_Chiroptera | Chiroptera species richness | Biodiversity mapping (Jenkins et al., 2013) | 10km |
| Hosts | Richness_10km_MAMMALS_mar2018_EckertIV_Eulipotyphla | Eulipotyphla species richness | Biodiversity mapping (Jenkins et al., 2013) | 10km |
| Hosts | Mammals/Richness_10km_MAMMALS_mar2018_EckertIV_Rodentia | Rodentia species richness | Biodiversity mapping (Jenkins et al., 2013) | 10km |
| Hosts | Richness_10km_MAMMALS_mar2018_EckertIV | Mammals species richness | Biodiversity mapping (Jenkins et al., 2013) | 10km |

References

Ballabio, C., P. Panagos, and L. Monatanarella, 2016: Mapping topsoil physical properties at European scale using the LUCAS database. *Geoderma* **261**, 110–123, DOI: 10.1016/j.geoderma.2015.07.006.

Buchhorn, M., B. Smets, L. Bertels, B.D. Roo, M. Lesiv, N.-E. Tsendbazar, M. Herold, and S. Fritz, 2020 (8. September): Copernicus Global Land Service: Land Cover 100m: collection 3: epoch 2018: Globe. . Zenodo.

Fick, S.E., and R.J. Hijmans, 2017: WorldClim 2: new 1-km spatial resolution climate surfaces for global land areas. *Int. J. Climatol.* **37**, 4302–4315, DOI: 10.1002/joc.5086.

Forsythe, W.C., E.J. Rykiel, R.S. Stahl, H. Wu, and R.M. Schoolfield, 1995: A model comparison for daylength as a function of latitude and day of year. *Ecol. Model.* **80**, 87–95, DOI: 10.1016/0304-3800(94)00034-F.

Jenkins, C.N., S.L. Pimm, and L.N. Joppa, 2013: Global patterns of terrestrial vertebrate diversity and conservation. *Proc. Natl. Acad. Sci.* **110**, E2602–E2610, DOI: 10.1073/pnas.1302251110.

Lebert, I., S. Bord, C. Saint-Andrieux, E. Cassar, P. Gasqui, F. Beugnet, K. Chalvet-Monfray, S.O. Vanwambeke, G. Vourc’h, and M. René-Martellet, 2022: Habitat suitability map of *Ixodes ricinus* tick in France using multi-criteria analysis. *Geospatial Health* **17**, DOI: 10.4081/gh.2022.1058.

Mu, H., X. Li, Y. Wen, J. Huang, P. Du, W. Su, S. Miao, and M. Geng, 2022: A global record of annual terrestrial Human Footprint dataset from 2000 to 2018. *Sci. Data* **9**, 176, DOI: 10.1038/s41597-022-01284-8.

Orgiazzi, A., R.D. Bardgett, and E. Barrios, 2016: Global Soil Biodiversity Atlas.

Robinson, T.P., G.R.W. Wint, G. Conchedda, T.P.V. Boeckel, V. Ercoli, E. Palamara, G. Cinardi, L. D’Aietti, S.I. Hay, and M. Gilbert, 2014: Mapping the Global Distribution of Livestock. *PLOS ONE* **9**, e96084, DOI: 10.1371/journal.pone.0096084.

Scharlemann, J.P.W., D. Benz, S.I. Hay, B.V. Purse, A.J. Tatem, G.R.W. Wint, and D.J. Rogers, 2008: Global Data for Ecology and Epidemiology: A Novel Algorithm for Temporal Fourier Processing MODIS Data. (Peter Gething, Ed.)*PLoS ONE* **3**, e1408, DOI: 10.1371/journal.pone.0001408.

Weiss, D., 2024 (24. June): A global map of travel time to cities. . DANS Data Station Physical and Technical Sciences.

Wint, W., D. Morley, and N.S. Alexander, 2013: Four Rodent and Vole Biodiversity Models for Europe. **1**, e3, DOI: 10.5334/jophd.ac.
